# The effect of residential aged care facility COVID-19 lockdowns on resident mortality

**DOI:** 10.1101/2023.06.26.23291907

**Authors:** Gabriela Uribe, Penelope Fotheringham, Corey Moore

## Abstract

**Aims:** This perspective is exploring the effect of residential aged care facility COVID-19 lockdowns on resident mortality.

**Relevance:** People living in residential aged care facilities (RACFs) are at increased risk of COVID-19 infection and death.

In Australia, COVID-19 infections in RACF residents represented seven percent of recorded infections and the case fatality rate (CFR) was 33%. RACFs were categorised as high-risk settings for COVID-19 and a number of measures to reduce the risk of acquiring COVID-19 were implemented at the National and State and Territory levels, such as strict infection control, monitoring, testing, vaccination and increased clinical capacity

**Methods:** All COVID-19 infections and deaths in the 12-months from 15 October 2021 to 14 October 2022 for the cohort of residents living in an Australian RACF (except Western Australia) at the beginning of this period were analysed.

**Key points:**

- Lockdowns of RACFs were successful in substantially reducing the number of COVID-19 infections and deaths in RACF residents. This was largely because the infection and mortality rates of COVID-19 were high in 2020 and 2021.
- The findings of this study could be used to evaluate and inform national and local states and territories’ guidelines on the management of COVID-19 in RACFs.

---

People living in residential aged care facilities (RACFs) are at increased risk of COVID-19 infection and death (1). Prior to the publication of specific Australian guidelines for COVID-19 management in March 2020, the attack rate (infection rate) in RACFs was reported at 64% (2) and 78% (3). In Australia, COVID-19 infections in RACF residents represented seven percent of recorded infections (4) and the case fatality rate (CFR) was 33% (5). RACFs were categorised as high-risk settings for COVID-19 and a number of measures to reduce the risk of acquiring COVID-19 were implemented at the National and State and Territory levels, such as strict infection control, monitoring, testing, vaccination and increased clinical capacity (4, 6, 7). In addition, lockdowns of entire facilities were enforced when a resident tested positive for COVID-19 (7). Australia also implemented strict public health measures on the entire community, including national and state lockdowns, to prevent community cases of COVID-19. This resulted in 100 times less deaths per capita in the aged care residents compared to the UK (8). These community level restrictions began to ease from October 2021.

For this study we analysed all COVID-19 infections and deaths in the 12-months from 15 October 2021 to 14 October 2022 for the cohort of residents living in an Australian RACF (except Western Australia) at the beginning of this period (9). Residents in RACFs located in Western Australia were excluded because of extended community isolation measures until March 2022. As publicly available aggregated data was used, ethics approval was not required.

There were 2,531 COVID-19 related deaths and 49,236 non-COVID-19 related deaths in the cohort of 173,305 residents from 15 October 2021 to 14 October 2022 (Figure 1). During this same 12-month period, RACFs were in lockdown for an average of 60 days, or 16.4% of the time. This corresponded to 23,341 resident-years of time in lockdown for this cohort. During this period, the CFR peaked at 18.8% before declining and then remaining between 2 and 5% (Figure 2).

**Figure 1.**
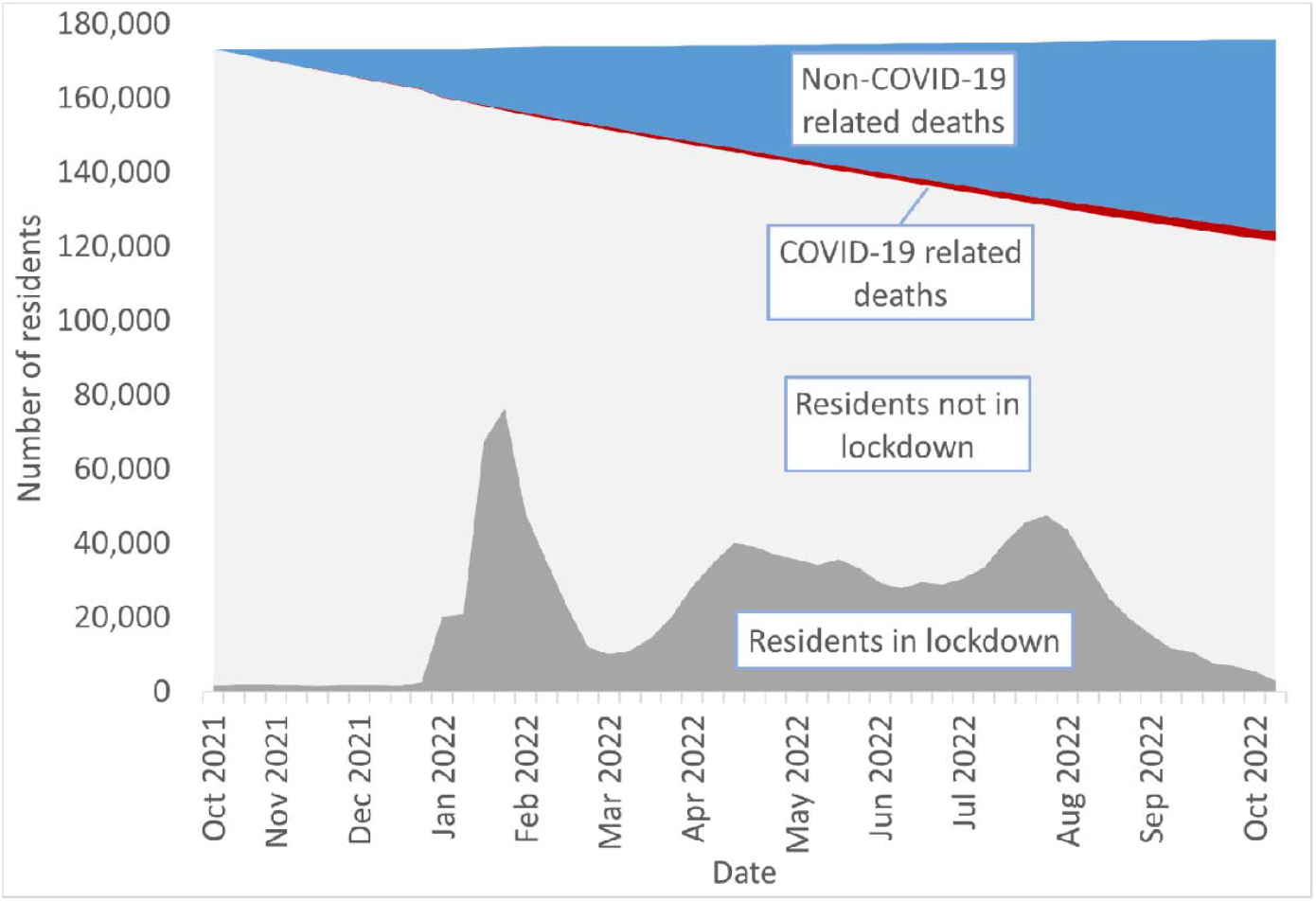
Non COVID-19, COVID-19-related death rates, number of residents in lockdown and in no lockdown for the period October 2021 – October 2022. Data source: GEN. GEN data: Providers, services and places in aged care (9)

**Figure 2.**
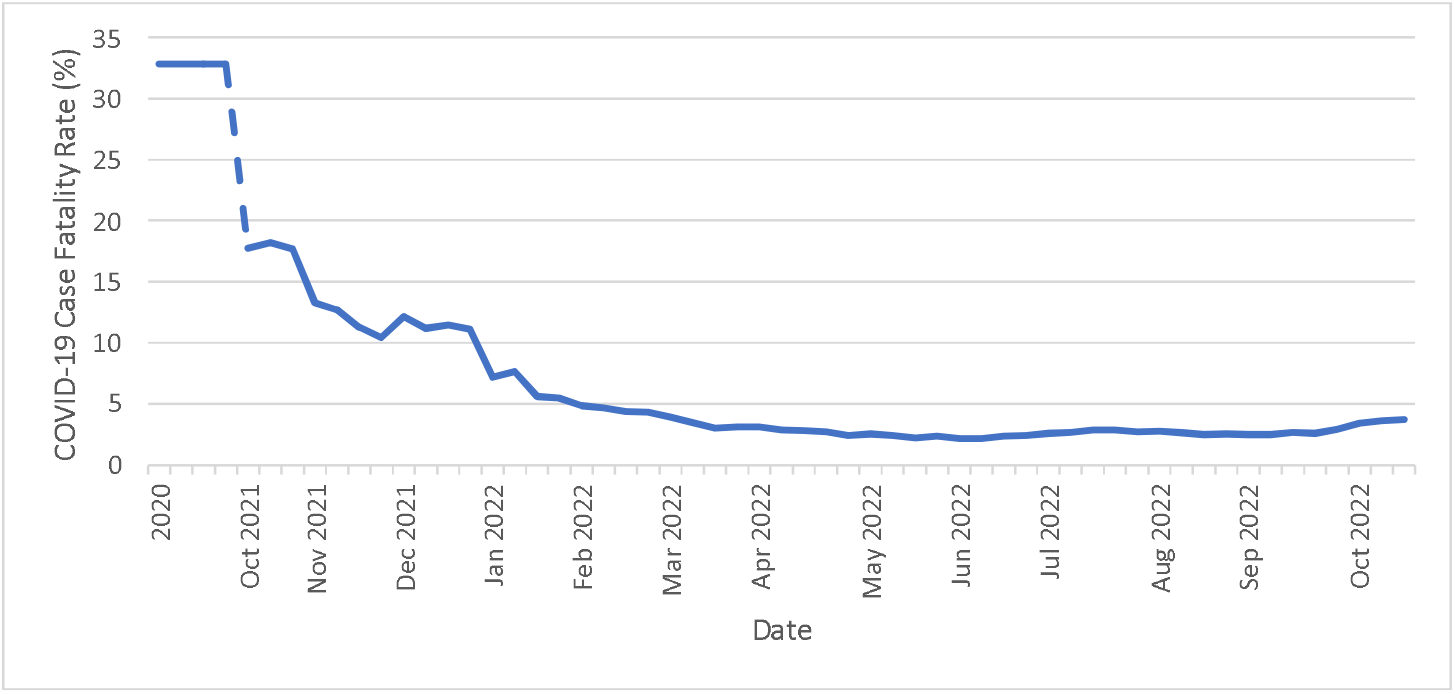
Case Fatality Rates in New South Wales for the Year 2020 to 2022. Data source: GEN. GEN data: Providers, services and places in aged care (9)

We then calculated that without lockdowns, there would have been an additional 18,421 years of life lost, based on a 70% attack rate and 18.8% CFR. Therefore, we estimate that, at a population level, for each day a resident was in lockdown, 0.8 days of life was saved. However, if we were to assume the current mortality rate of 5% and the same pattern of lockdowns going forward, we estimate that one day in lockdown would result in 0.2 days of life saved.

Lockdowns of RACFs were successful in substantially reducing the number of COVID-19 infections and deaths in RACF residents. This was largely because the infection and mortality rates of COVID-19 were high in 2020 and 2021. These rates have since decreased, possibly due to natural attenuation of the virus, acquired immunity, vaccinations, antivirals, improved clinical management, and the survival effect. The findings of this study, together with other factors such as the predicted burden on the healthcare system, community attitudes, and impact of isolation in residents’ health and well-being (1, 10), could be used to evaluate and inform national and local states and territories’ guidelines on the management of COVID-19 in RACFs.

## Supporting information

Assumptions made for the modelling

## Data Availability

All data used in this manuscript is available online at https://www.gen-agedcaredata.gov.au/Resources/Access-data/2023/April/GEN-data-Providers,-services-and-places-in-aged-ca

## Acknowledgements

Not applicable.

