## Supplementary material for "The effect of residential aged care facility COVID-19 lockdowns on resident mortality": Assumptions made for the modelling

**Supplementary File. Assumptions made for the modelling.**

- All cases and deaths were reported in the week they occurred and were correct at the time.
- Where there was no reported data from an RACF during an outbreak, the case and death numbers were interpolated.
- From 23 September 2022, RACF case and death numbers of 5 or less were reported as “<6”. These were assumed to be 2.
- An attack rate of 70%, based on reports in early 2020 (1, 2).
- The number of RACFs was calculated to be 2,464 (excluding WA), with 173,305 residents (3).
- All residents of a RACF were considered to be in lockdown when the RACF was in lockdown.
- COVID-19 deaths and infections were randomly distributed among residents.
- Deaths not due to COVID-19 were modelled using exponential decay with a medium age of 36 months (3).
- Reinfections were not considered.
- For Figure 2, a single CFR was calculated for each outbreak and then combined to form the weekly national average.
- For the final model, the attack rate was left unchanged at 70%. The justification being: 1. to reduce the number of assumptions, model complexity, and the need for a sensitivity analysis; 2. a different attack rate would affect both the number of infections as well as deaths; and 3. the outcome of decreased effectiveness of lockdowns, would be the same even with a different attack rate.
